# Generate Synthetic Data in R for a Hypothetical Alzheimer’s Disease Trial

**DOI:** 10.1101/2024.02.05.24302140

**Authors:** Ron Handels, Linus Jönsson, Lars Lau Raket, the Alzheimer’s Disease Neuroimaging Initiative

## Abstract

**INTRODUCTION:** Representative data of recent Alzheimer’s Disease (AD) trials are difficult to obtain. We aimed to generate a synthetic version of an original real-world observational dataset, subsequently apply a plausible AD treatment effect, and make our method open-source available.

**METHODS:** Synthetic data was generated in the following steps: (1) Obtain real-world data from the ADNI study on demographic (age, sex, education), clinical (cognition: MMSE and ADAS; function: FAQ; composite cognition/function: CDR, ADCOMS) and biological (genetics: APOE4; cerebrospinal fluid: ABeta, Tau; imaging: PET-SUVR-centiloid) outcomes at baseline, 6, 12 and/or 18-month follow-up (35 variables), with missing data multiple-imputed to obtain 10 sets of 537 individuals. (2) Estimate (theoretical) minimum and maximum (all continuous variables) and proportions (all categorical variables). (3) Rescale to 0-1 range (continuous). (4) Estimate beta distribution shape parameters (method of moments; continuous). (5) Transform to cumulative probability distribution function (using shape parameters; continuous) and to cumulative probability (categorical). (6) Transform to a normal distribution. (7) Estimate variance-covariance matrix. (8) Generate random correlated normal data using Cholesky decomposition of variance-covariance. (9) Transform to cumulative probability distribution function. (10) Transform to beta distribution (using shape parameters; continuous). (11) Rescale to original range. (12) Keep half as control arm, and half as intervention arm, and estimate change from baseline. (13) Multiply intervention change from baseline with self-defined hypothetical relative treatment effect. We assumed correlations on normalized scale were similar to correlations on original scale. R code is available on github: https://github.com/ronhandels/synthetic-correlated-data.

**RESULTS:** The synthetic distribution and mean over time showed large similarity to the original data (visually assessed). The absolute difference in pairwise correlations between original and synthetic data median was 0.02 (95th percentile=0.11, max=0.18).

**CONCLUSION:** We judged our method sufficiently valid to generate synthetic correlated plausible hypothetical trial results.

## Introduction

Recent Alzheimer’s disease (AD) treatments have shown significant effects on short-term surrogate outcomes [van Dyck et al, 2023; Sims et al, 2023]. To assess the health-economic impact of AD treatment their short-term outcomes must likely be extrapolated over lifetime using a decision-analytic model. To improve understanding, transparency and credibility of different available models they have been cross-compared using a conceptually defined benchmark treatment effect [Handels et al., 2023]. This could be improved by using real-world individual-level trial data. However, we experienced limited availability of data from AD trials.

Individual-level data in healthcare are controlled due to privacy issues and therewith limitedly available. A recent review has showed 7 use cases of synthetic datasets in health care, being “a) simulation and prediction research, b) hypothesis, methods, and algorithm testing, c) epidemiology/public health research, d) health IT development, e) education and training, f) public release of datasets, and g) linking data” [Gonzales et al., 2023].

We aimed to generate a synthetic version of an original real-world observational dataset, subsequently apply a plausible AD treatment effect, and make our method open-source available. The results of this study are targeted for being implemented in health-economic decision-analytic models and subsequently cross-compare them on health-economic outcomes.

## Methods

We (RH, LJ, LLR) determined the characteristics of a plausible future trial based on an open discussion. This was set in terms of participants recruited in a memory clinic with the following inclusion criteria: age between 55-85 years; objective impairment in episodic memory (based on Wechsler Memory Scale-IV Logical Memory II); amyloid positive via amyloid cerebrospinal fluid Amyloid Beta or amyloid Positron Emission Tomography (PET); Clinical Dementia Rating (CDR) = 0.5; Mini-Mental State Examination (MMSE) ≥ 24; and the following exclusion criteria: MRI based confounding pathologies (e.g., acute or sub-acute hemorrhage); Unstable dose of Acetylcholinesterase inhibitor (AChEI).

We considered a sub-selection of the data from the ADNI as representative for this target population. Data from the ADNI has been collected in a multicenter longitudinal study the US and Canada and tracked AD progression with clinical, imaging, genetic and biospecimen biomarkers through the process of normal aging, mild cognitive impairment and dementia.

Data used in the preparation of this article were obtained from the Alzheimer’s Disease Neuroimaging Initiative (ADNI) database (adni.loni.usc.edu). The ADNI was launched in 2003 as a public-private partnership, led by Principal Investigator Michael W. Weiner, MD. The primary goal of ADNI has been to test whether serial magnetic resonance imaging (MRI), positron emission tomography (PET), other biological markers, and clinical and neuropsychological assessment can be combined to measure the progression of mild cognitive impairment (MCI) and early Alzheimer’s disease (AD).

We considered that the following selection of ADNI participants reflected this target setting and selection criteria. Therefore, we estimated the distribution, mean over time (except what we determined as demographics only obtained at baseline) and correlation of a set of variables. The following set of variables were expected to be measured in a plausible future trial and available in ADNI: age, sex, years of education, ethnicity Hispanic or Latino yes/no, ApoE4 status, CDR sum of boxes (CDR-SB), CDR global score, MMSE total score, Alzheimer’s Disease Assessment Scale-cognitive subscale (ADAS-Cog13) total score, Functional Activities Questionnaire (FAQ) total score, AD Composite Score (ADCOM), cerebrospinal fluid (CSF) amyloid beta (ABETA), CSF total tau (TAU), CSF phosphorelated tau (PTAU), Amyloid PET standardised uptake value ratios centiloid score (PET-SUVr-CL) based on PIB, AV45 or Florbetaben.

### Box 1

**steps for generating synthetic data**

#### Part A: Original Data

1. Original real-world data from the Alzheimer’s Disease Neuroimaging Initiative (ADNI) study on
  i. demographic (age, sex, education),
  ii. clinical (cognition: Mini-Mental State Examination (MMSE) and Alzheimer’s Disease Assessment Scale (ADAS); function: Functional Activities Questionnaire (FAQ); composite cognition/function: Clinical Dementia Rating (CDR), Alzheimer’s Disease Composite Score (ADCOMS)) and
  iii. biological (genetics: APOE4; cerebrospinal fluid: ABeta, Tau; imaging: PET-SUVR-centiloid) outcomes at baseline, 6, 12 and/or 18-month follow-up (35 variables), with missing data multiple-imputed to obtain 10 sets of 537 individuals.
2. Estimate (theoretical) minimum and maximum (all continuous variables) and proportions (all categorical variables).
3. Rescale to 0-1 range (continuous).
4. Estimate beta distribution shape parameters (method of moments; continuous).
5. Transform to cumulative probability distribution function (CDF) (using shape parameters; continuous) and to cumulative probability (categorical).
6. Transform to a normal distribution (using quantile function, i.e., inverse cumulative distribution function).
7. Estimate variance-covariance matrix.

#### Part B: Synthetic data

8. Generate random correlated normal data (mean=0, SD=1) using Cholesky decomposition of variance-covariance matrix from step 7.
9. Transform to cumulative probability distribution function (CDF).
10. Transform to beta distribution (using quantile function, i.e., inverse cumulative distribution function) (using beta distribution shape parameters from step 4; continuous).
11. Rescale to original range (using minimum and maximum and proportions from step 2).

#### Part C: Treatment effect

12. Keep half as control arm, and half as intervention arm, and estimate change from baseline.
13. Multiply intervention change from baseline with self-defined hypothetical relative treatment effect.

Data from the merged ADNI file were used as a basis. Data from additional files on CDR, ADAS and MMSE were added to obtain item-level scores to calculate the ADCOMS score. Data from additional file florbetaben were added to obtain PET SUVR florbetaben scores. Multiple imputation was used to impute missing data.

### Generating synthetic data

The selected data from the ADNI was synthetically recreated (part A and B described in box 1 as steps 1-11, also visualized in Figure 1 with matching step numbers and colors). Then, a hypothetical treatment effect was implemented by manually manipulating half of the synthetic (part C described in box 1 as steps 12-13).

**Figure 1:**
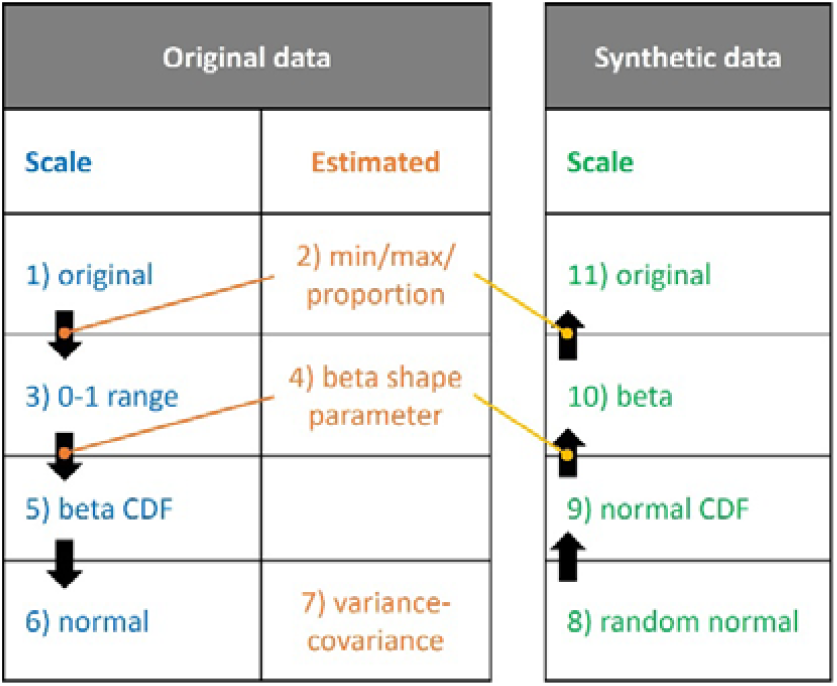
Overview of steps (1-11) taken to create synthetic correlated data from original data (abbreviation: CDF, cumulative probability distribution function).

### Validating synthetic data

Original ADNI data and its synthetically recreated data were plotted in terms of distribution, mean over time and correlation. The validity of the synthetic data regarding these 3 aspects was visually assessed.

### R code and tutorial

A simplified version of the code including a tutorial of the validation for part A and B (steps 1-11) is available on GitHub via https://github.com/ronhandels/synthetic-correlated-data. An ISPOR conference poster describing our method is also available [Handels et al., 2023].

We note the example data available on GitHub was generated by taking the original ADNI data and adding a fixed change as well as large random variation to each variable. Therefore, this example data does not contain any original data from ADNI data, it does not represent ADNI data, and we think it does not represent clinical correctness. We therefore recommend using the example data only for the purpose of understanding our method in our tutorial in which these example data are used.

## Results

The distribution and mean over time from the synthetic data showed relatively large similarity to the original data (visually assessed, see Figure 2 and Figure 3). The absolute differences in pairwise correlations between original data and synthetic data showed had a median of 0.04, a 95th percentile of 0.20 and a maximum of 0.30. Figure 4 shows a heatmap of each absolute difference in pairwise correlation.

**Figure 2:**
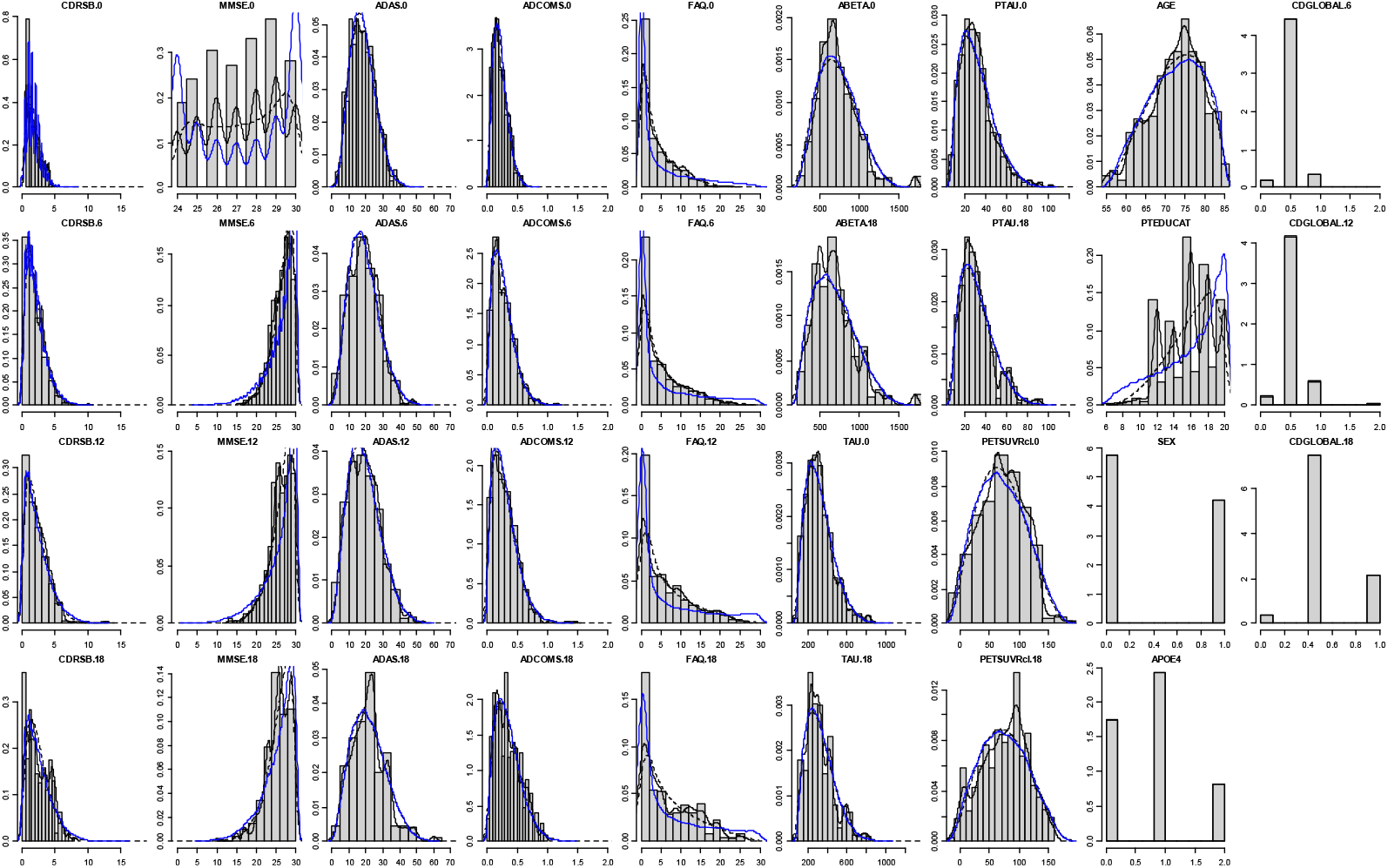
Histogram of original data, density line of original data (black solid line), density line of the beta distribution parameters (black dashed line), and density line of simulated data (blue) all on rescaled 0-1 range. Density line of original data should be similar to density line of the beta distribution parameters as well as to the density line of the simulated data.

**Figure 3:**
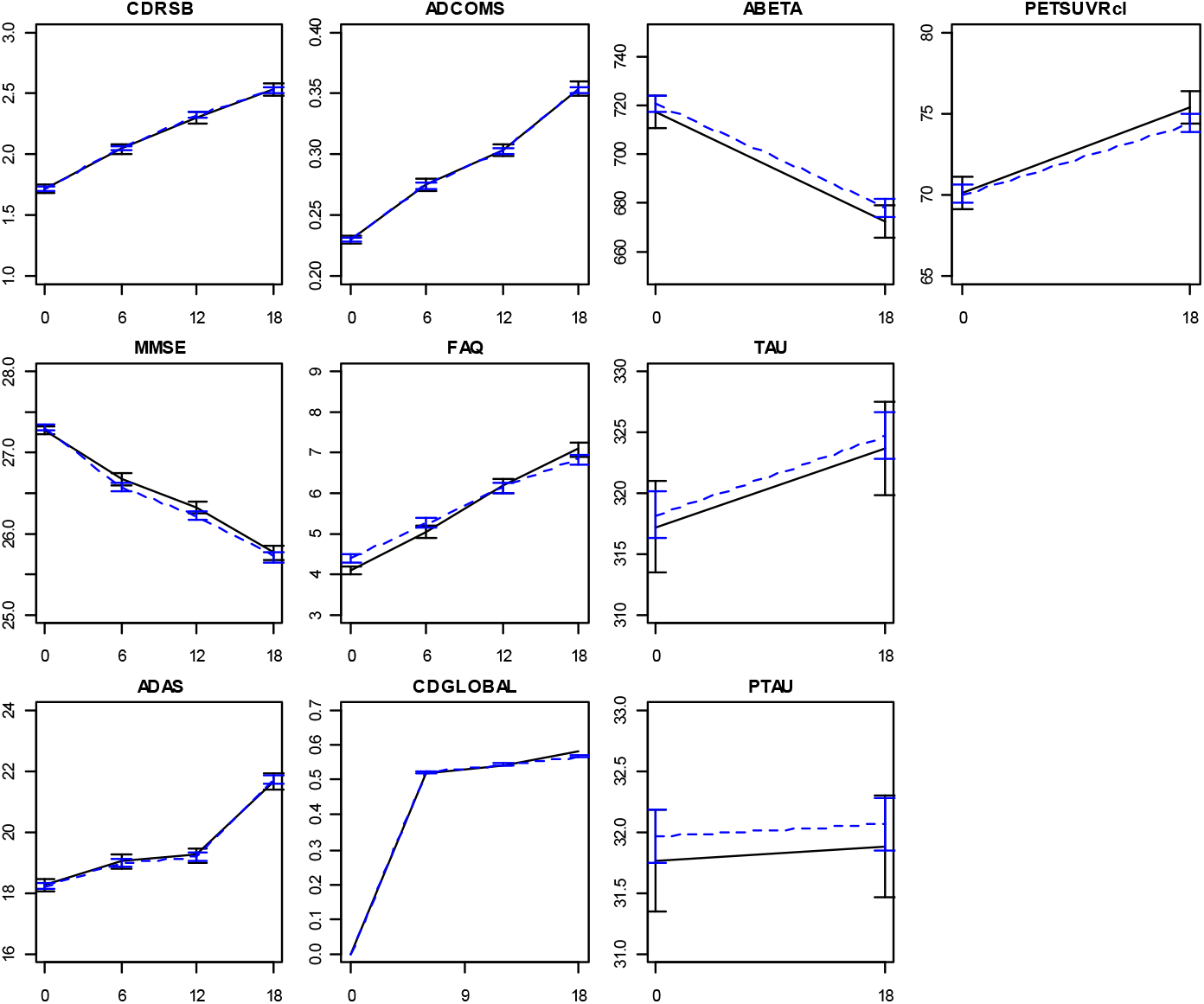
Mean and 95% confidence interval of original data and synthetically recreated data over time.

**Figure 4:**
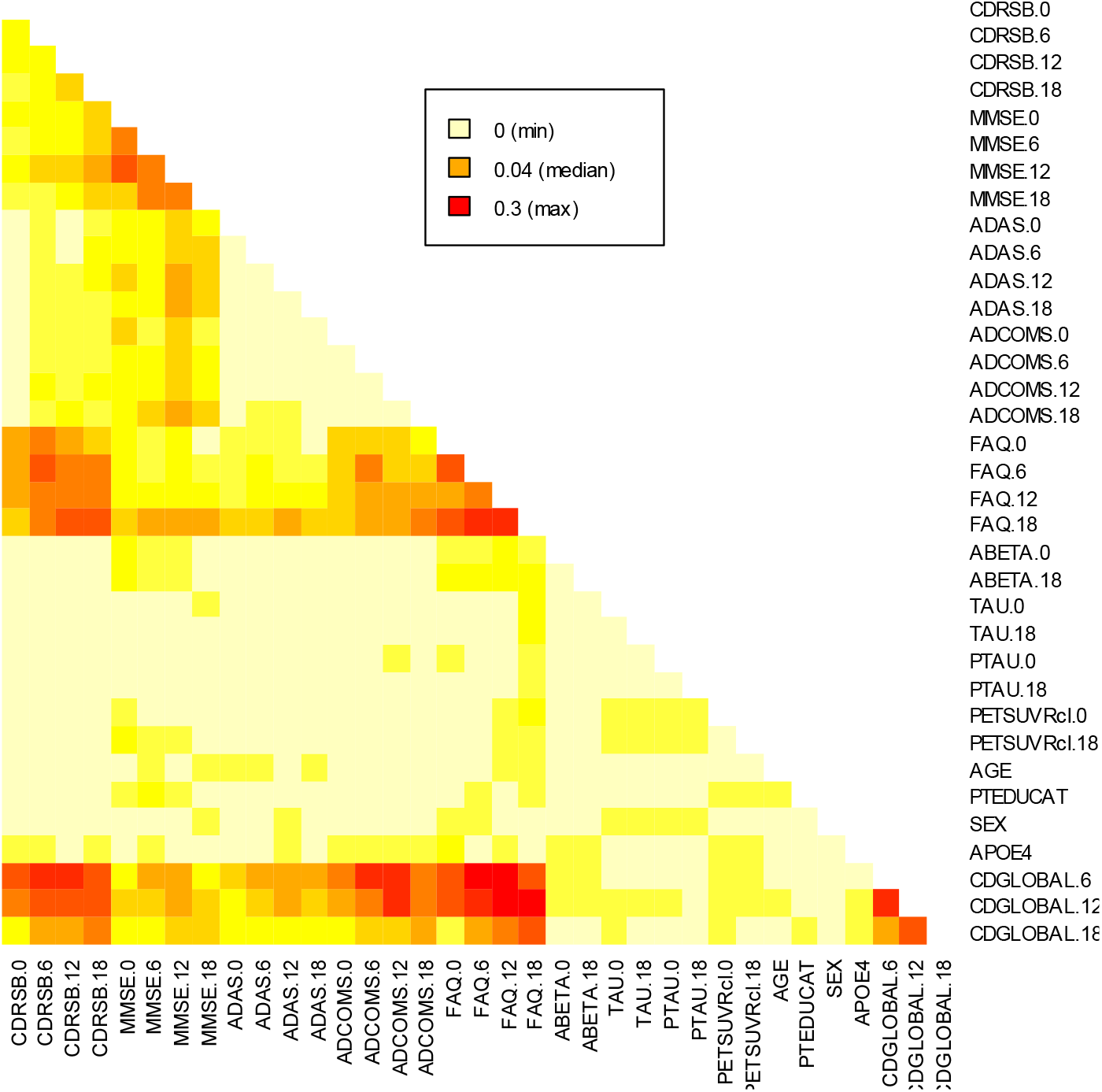
Heatmap of the absolute values of the correlation matrix fitted to the original data (on its original scale) minus the correlation matrix fitted to the simulated data (on its final scale).

After applying the self-defined hypothetical relative treatment effect, the data were summarized into a set of tables and graphs which we considered plausible in terms of being reported in a real-world trial publication. These can be found in Tables 1, Table 2 and Table 3, and Figure 5. These include reportion of confidence intervals and results of regression analysis to test for significant differences.

**Table 1:** Trial hypothetical inclusion and exclusion criteria.

|  |
| --- |
| **Inclusion criteria:**<br>Age between 55-85 years<br>Objective impairment in episodic memory (based on Wechsler Memory Scale-IV Logical Memory II)<br>Amyloid positive* via amyloid cerebrospinal fluid Amyloid Beta or amyloid PET<br>CDR = 0.5<br>MMSE ≥ 24 |
| **Exclusion criteria:**<br>MRI based confounding pathologies (e.g., acute or sub-acute hemorrhage)<br>Unstable dose of AChEI |

\* Definitions of positivity might vary across studies/measures/tools.

**Table 2:** Sample characteristics at baseline (standard deviation of the mean).

|  | Control (SOC + placebo)<br>n=654 | Intervention (SOC + DMT)<br>n=654 | p-value |
| --- | --- | --- | --- |
| Age, mean (SD) | 73 (6.7) | 73 (6.8) | 0.420 |
| Female, % | 42% | 39% | 0.400 |
| Education (years), mean (SD) | 16.5 (3.5) | 16.1 (3.6) | 0.079 |
| ApoE e4/e4, % | 18% | 19% | 0.620 |
| CDR SOB, mean (SD) | 1.71 (1.01) | 1.70 (0.99) | 0.870 |
| MMSE, mean (SD) | 27.4 (2.4) | 27.2 (2.4) | 0.350 |
| ADAS-Cog13, mean (SD) | 18.0 (7.4) | 18.6 (7.3) | 0.110 |
| ADCOMS, mean (SD) | 0.23 (0.12) | 0.23 (0.12) | 0.580 |
| FAQ, mean (SD) | 4.30 (6.72) | 4.43 (6.73) | 0.730 |
| CSF ABeta24 (pg/mL), mean (SD) | 718 (242) | 705 (232) | 0.340 |
| CSF total tau (pg/mL), mean (SD) | 311 (133) | 325 (146) | 0.067 |
| CSF phosphorylated tau (pg/mL), mean (SD) | 31 (15.0) | 33 (16.5) | 0.023 |
| Amyloid PET SUVR centiloid, mean (SD) | 68 (40) | 72 (42) | 0.041 |

Abbreviations: ADAS-Cog13, Alzheimer's Disease Assessment Scale-cognitive subscale; ADCOMS, AD Composite Score; CDR SOB, Clinical Dementia Rating sum of boxes; CSF, cerebrospinal fluid; DMT, disease-modifying treatment; FAQ, functional activities questionnaire; MMSE, Mini-Mental State Examination; PET SUVR, positron emission tomography standard uptake value ratio; SD, standard deviation; SOC, standard of care.

**Table 3:** Efficacy outcomes in terms of number or mean change from baseline after 18 months (standard error).

|  | Control (SOC + placebo)<br>n=654 | Intervention (SOC + DMT) n=654 | p-value |
| --- | --- | --- | --- |
| CDR global, n (%) |  |  | 0.463 |
| -0.5 (from 0.5 to 0) | 34 (5%) | 54 (8%) |  |
| +0 (from 0.5 to 0.5) | 484 (74%) | 473 (72%) |  |
| +0.5 (from 0.5 to 1) | 136 (21%) | 127 (19%) |  |
| +1.5 (from 0.5 to 2) | 0 | 0 |  |
| +2.5 (from 0.5 to 3) | 0 | 0 |  |
| CDR-SB, mean (SE) | 0.84 (0.06) | 0.59 (0.06) | 0.004 |
| MMSE, mean (SE) | -1.7 (0.2) | -1.1 (0.2) | 0.021 |
| ADAS-Cog13, mean (SE) | 3.8 (0.3) | 2.2 (0.3) | 0.000 |
| ADCOMS, mean (SE) | 0.12 (0.01) | 0.08 (0.01) | 0.000 |
| FAQ, mean (SE) | 2.6 (0.3) | 1.9 (0.3) | 0.058 |
| CSF ABeta24 (pg/mL), mean (SE) | -36 (6) | 267 (6) | 0.000 |
| CSF total tau (pg/mL), mean (SE) | 6.8 (2.3) | -125.7 (2.3) | 0.000 |
| CSF phosphorylated tau (pg/mL), mean (SE) | 0.2 (0.3) | -20.1 (0.3) | 0.000 |
| Amyloid PET SUVR centiloid, mean (SE) | 5.9 (0.9) | -45.3 (0.9) | 0.000 |

Abbreviations: ADAS-Cog13, Alzheimer's Disease Assessment Scale-cognitive subscale; ADCOMS, AD Composite Score; CDR SOB, Clinical Dementia Rating sum of boxes; CSF, cerebrospinal fluid; DMT, disease-modifying treatment; FAQ, functional activities questionnaire; MMSE, Mini-Mental State Examination; PET SUVR, positron emission tomography standard uptake value ratio; SE, standard error; SOC, standard of care.
The data reflected an intention-to-treat population and were assumed adjusted for conditional missingness as this was to be expected in practice.

**Figure 5:**
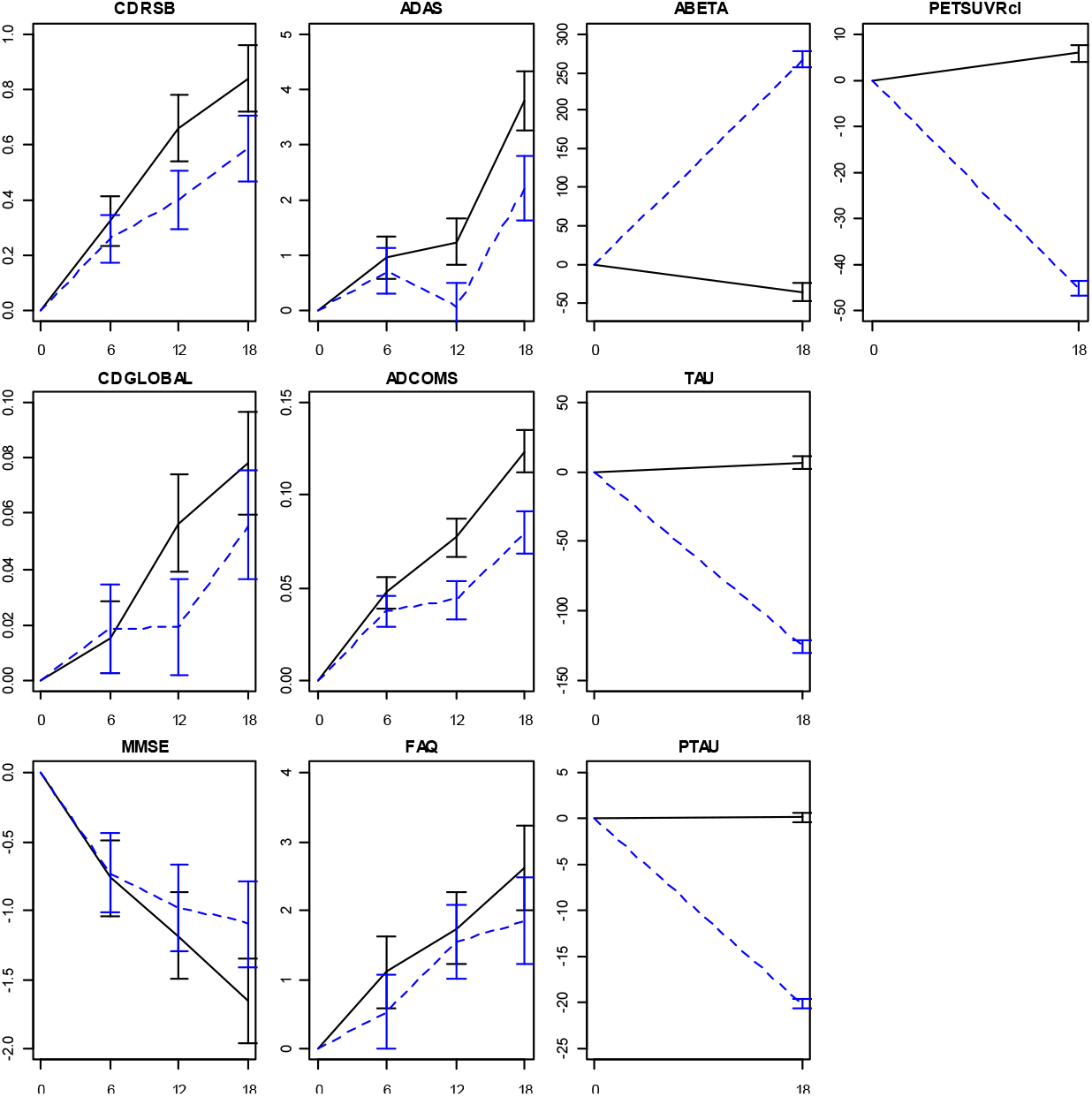
Efficacy outcomes: mean change from baseline (standard error presented in bars).

## Discussion

We generated plausible AD treatment effect individual-level data by synthetically recreating real-world observational dataset combined with a hypothetical treatment effect.

We think the similarity between the original and synthetically recreated data argues for their face validity, and therewith its usability as benchmark for cross-comparing health-economic decision-analytic models in AD.

We note the self-defined hypothetical relative effect was not disclosed as the hypothetical trial results have been aimed for being implemented in health-economic decision-analytic models. For the purpose of implementing them the original intended treatment effect is not disclosed.

Among alternative methods for generating synthetic correlated data from different distributions is R package ‘synthpop’ [Nowok et al., 2016]. A comparison to this method is beyond the scope of our study, as is a review of alternative technologies from areas of multiple imputation, generalized mixed models or artificial intelligence.

### Limitations

The results of this study are subject to several limitations. First, correlations among the data were estimated on normalized scale and assumed representative for correlations on the original scale. This assumption is likely incorrect in case data is not normally distributed.

However, for much data the impact seems relatively small as reflected by a relatively small deviation on correlation in the observed and synthetically recreated data. Second, missing data or drop-out were not simulated, limiting the representativeness of the data to a real-world setting in which missing data or drop-out in trials is common. Third, generally we believe synthetic data are as good as the underlying models parameterizing them. Our method is based on the assumption that the data can be correctly described by the parameters of the beta distribution and the correlation coefficient. Likely, data from multimodal distribution or with non-linear associations are incorrectly synthetically recreated by our method.

## Conclusion

We generated a synthetic version of an original real-world observational dataset, applied a plausible AD treatment effect, and made our method open-source available. We judged our method sufficiently valid to generate synthetic correlated plausible hypothetical trial results.

## Data Availability

Part of the data are available online at https://github.com/ronhandels/synthetic-correlated-data.
All data produced in the present study are available upon reasonable request to the authors.

## Acknowledgment

Data collection and sharing for this project was funded by the Alzheimer’s Disease Neuroimaging Initiative (ADNI) (National Institutes of Health Grant U01 AG024904) and DOD ADNI (Department of Defense award number W81XWH-12-2-0012). ADNI is funded by the National Institute on Aging, the National Institute of Biomedical Imaging and Bioengineering, and through generous contributions from the following: AbbVie, Alzheimer’s Association; Alzheimer’s Drug Discovery Foundation; Araclon Biotech; BioClinica, Inc.; Biogen; Bristol-Myers Squibb Company; CereSpir, Inc.; Cogstate; Eisai Inc.; Elan Pharmaceuticals, Inc.; Eli Lilly and Company; EuroImmun; F. Hoffmann-La Roche Ltd and its affiliated company Genentech, Inc.; Fujirebio; GE Healthcare; IXICO Ltd.; Janssen Alzheimer Immunotherapy Research & Development, LLC.; Johnson & Johnson Pharmaceutical Research & Development LLC.; Lumosity; Lundbeck; Merck & Co., Inc.; Meso Scale Diagnostics, LLC.; NeuroRx Research; Neurotrack Technologies; Novartis Pharmaceuticals Corporation; Pfizer Inc.; Piramal Imaging; Servier; Takeda Pharmaceutical Company; and Transition Therapeutics. The Canadian Institutes of Health Research is providing funds to support ADNI clinical sites in Canada. Private sector contributions are facilitated by the Foundation for the National Institutes of Health (www.fnih.org). The grantee organization is the Northern California Institute for Research and Education, and the study is coordinated by the Alzheimer’s Therapeutic Research Institute at the University of Southern California. ADNI data are disseminated by the Laboratory for Neuro Imaging at the University of Southern California.

## Conflict of interest

LLR is an employee of Eli Lilly and Company.

RH received outside this study research grants from JPND, ZonMW, IMI, H2020 (paid to institution); received outside this study consulting fees in the past 3 years from Lilly Nederland (2023), iMTA (2023), and Biogen (2021) (paid to institution); is member of IPECAD and member of ISPOR special interest group open-source models (un-paid).

## Funding

This study did not receive any funding.

## Notes

### Clinical Protocols

https://github.com/ronhandels/synthetic-correlated-data

### Author Declarations

We requested the data from the Alzheimer's Disease Neuroimaging Initiative (ADNI) for the following specific aim: "describe the natural progression over a short-term period by mimicing/emulating data typically obtained from AD drug treatment randomized trials" and method as described in our manuscript (in short: select data from the ADNI, fit variance-covariance matrix, use variance-covariance matrix to generate synthetic data (mimic/emulate the data)). We have received the following reply from ADNI: "Your request for access to the Alzheimer's Disease Neuroimaging Initiative (ADNI) Data has been approved."

